# The Effectiveness of Telephone Intervention for Improving Patient Adherence to Medication among Diabetic Patients: A Systematic Review and Meta-analysis of Randomized Trials

**DOI:** 10.1101/2021.06.20.21259200

**Authors:** Debbie P. Monterona, Rhoda Alfonsa Matinong, Jeriel De Silos

**Affiliations:** Department of Family and Community Medicine, De La Salle Health and Sciences Institute

## Abstract

**Introduction:** Diabetes is one of the chronic diseases that requires adherence to prescribed medications. With the current pandemic, mobile technology plays a role in caring for patients remotely.

**Objective:** To determine the effectiveness of telephone intervention (phone call and text message) on medication adherence among diabetic patients.

**Methodology:** Randomized controlled trials were searched in Cochrane Library, PubMed, Herdin, BMC Health Services Research using combination of terms through boolean operators (“phone message” OR “phone call” AND (“medication adherence” AND “diabetes”) which compared telephone intervention vs usual care. mean, sample size and standard deviation of Medication Adherence in each study were extracted. Review Manager 5.4 software was used for statistical analysis.

**Results:** Three trials met the inclusion criteria and were included in this study. The telephone intervention did not result in statistically significant improvement in medication adherence among diabetics (pooled mean difference: 0.05 95%CI -.08 to 0.17) *Conclusion:* The intervention was no more effective than the usual care. However, mobile use has potential application for remote care during this pandemic.

## INTRODUCTION

Diabetes is a major global health problem. It is a chronic, metabolic disease characterized by elevated levels of blood sugar which may cause damage to the heart, blood vessels, eyes, kidneys and nerves. According to the World Health Organization (WHO), between the years 2000 and 2019 there was 70% increase in deaths from diabetes globally, with an 80% rise in deaths among males. (1)

Individuals with diabetes may not have enough insulin produced in the body or does not respond properly to insulin. The common types of diabetes are: Type 1 Diabetes, Type 2 Diabetes and Gestational Diabetes. In Type 1 Diabetes Mellitus, which usually occurs in childhood or early adulthood, the immune system destroys the cells of the pancreas that make insulin. Type 2 Diabetes Mellitus is caused by the body’s ineffectiveness to use insulin properly. This type often results from lack of physical activity and obesity. The third type which occurs during pregnancy is Gestational Diabetes. (2) Controlling blood sugar is very important to prevent serious complications. There are so many ways to control blood sugar: diet therapy, regular exercise, weight control, cessation of smoking, maintaining normal blood pressure and medical therapy.(3)

Chronic diseases are the major cause of death and disability worldwide. According to data from the World Health Organization, in the Philippines, chronic diseases accounted for 57% of all deaths in 2002. In that year alone, the total deaths in the Philippines were 449,000 and 253,000 deaths were due to chronic diseases: Cardiovascular disease 27%, Cancer 9%, Chronic Respiratory Disease 6%, Diabetes 3% and other chronic diseases 12%.(4)

Medication non-adherence is a growing concern globally. In terms of managing chronic diseases like hypertension and diabetes, long-term adherence to medications is a vital component.

Adherence is defined by WHO as “The extent to which a person’ s behavior— taking medication, following a diet, or making healthy lifestyle changes—corresponds with agreed-upon recommendations from a health-care provider.”(5) Medication adherence can be assessed using direct or indirect methods. Direct methods include drug detection in biological fluid and directly observing intake of medication. Indirect methods are commonly used and may include interviews, surveys, pill counts and refill records.(6) Examples of these methods are The 8-Item Morisky Medication Adherence Scale, a self-report validated assessment tool that measures non-adherence (7) and Medication Possession Ratio that is used to measure adherence thru refill records. The non-adherence can result to exacerbation of the chronic condition, hospitalization and even death.

Various methods can be implemented to improve non-adherence such as simplifying medication regimen, explaining key information when prescribing, providing behavioral support especially for the elderly, scheduling follow-ups to monitor medication adherence and use medication adherence improving aids like medication calendars and charts.(8)

There has been a rapid growth in Mobile technology all around the globe for the past decade. In the Philippines, there were around 29.7M smartphone users for the year 2016. (9) Mobile telecommunication companies like Smart, Globe and SunCellular have various prepaid and postpaid plans giving customers unlimited text messages and calls. This technology has improved communication among healthcare providers. Abroad, phone call intervention is being used in healthcare system in following up patients that improves patient care, patient’s satisfaction and health care providers as well.(10)

Adherence to medications is integral to the management of Non-Communicable Diseases such as Diabetes. This is one of the most common types of chronic diseases seen in a primary care setting like the Family and Community Medicine outpatient clinic. Treatment algorithms are well in place for most, if not these chronic diseases. However, there are various factors that may affect treatment outcomes. In the context of a developing country like the Philippines, there are a multitude of reasons for failure to help patients with chronic diseases. A very vital factor to treating such diseases and oftentimes overlooked is the patients’ adherence to medications.

With the present Covid-19 pandemic, the use of telehealth has been found to be very useful in reaching out and helping the communities, the families and individuals with their health concerns in a remote setting.

There are randomized controlled trials conducted on the effect of phone interventions on medication adherence among patients with chronic diseases but no systemic reviews nor meta-analysis done on diabetic patients. This study aims to determine the effectiveness of telephone intervention (text message or phone call reminder) in helping diabetic patients adhere to their medications. Probably this can be used in the hospital or clinics as a way of helping the patients with diabetes to ensure that they take their medications which will lessen hospitalizations, emergency calls and other complications.

## METHODOLOGY

This meta-analysis review is guided by Cochrane Handbook for Systematic Reviews of Interventions and written in accord with the Preferred Reporting Items for Systematic Reviews and Meta-analysis (PRISMA).

Trials with the following criteria were included: 1) randomized clinical trials evaluating telephone intervention (text message or phone call reminder) to promote medication adherence in adults with diabetes. 2) Participants in this study are adult patients ≥18 years old and above with diagnosis of Diabetes, 2) at least prescribed with 1 oral hypoglycemic medication and 3) possessing a cell phone and knowing how to answer the telephone or have someone to assist with reading the text message

Exclusion criteria were as follows: 1) patients requiring emergency intervention or admission. 2) patients with mental illness or any other long-term health conditions such as HIV, Cancer, End stage Renal disease requiring hemodialysis 3) pregnant patients

Studies with medication adherence measured by Medication Possession Ratio, the 8 item Adherence Scale post-intervention became part of the study.

Electronic databases were searched which included PubMed, Herdin, Cochrane Library, and BMC Health Services Research. For the electronic search, combination of terms through boolean operators (“phone message” OR “phone call” AND (“medication adherence” AND “diabetes”). The bibliographies of included studies and any relevant systematic reviews identified were checked for further references to relevant trials.

Titles and abstracts published from January 2000-October 2020 were downloaded, screened and duplicates removed. The two reviewers read the full text articles and evaluated if to be included or not. Information extracted from each study included study design, country, age, population characteristics, sample size, intervention and the specific outcomes of interest.

Extraction of data from each study included the mean, sample size and standard deviation of Medication Adherence. Review Manager 5.4 software was used for statistical analysis.

The investigators assessed the risk of bias using Cochrane Handbook for Systematic reviews of Interventions as a guide.

**Figure 1.**
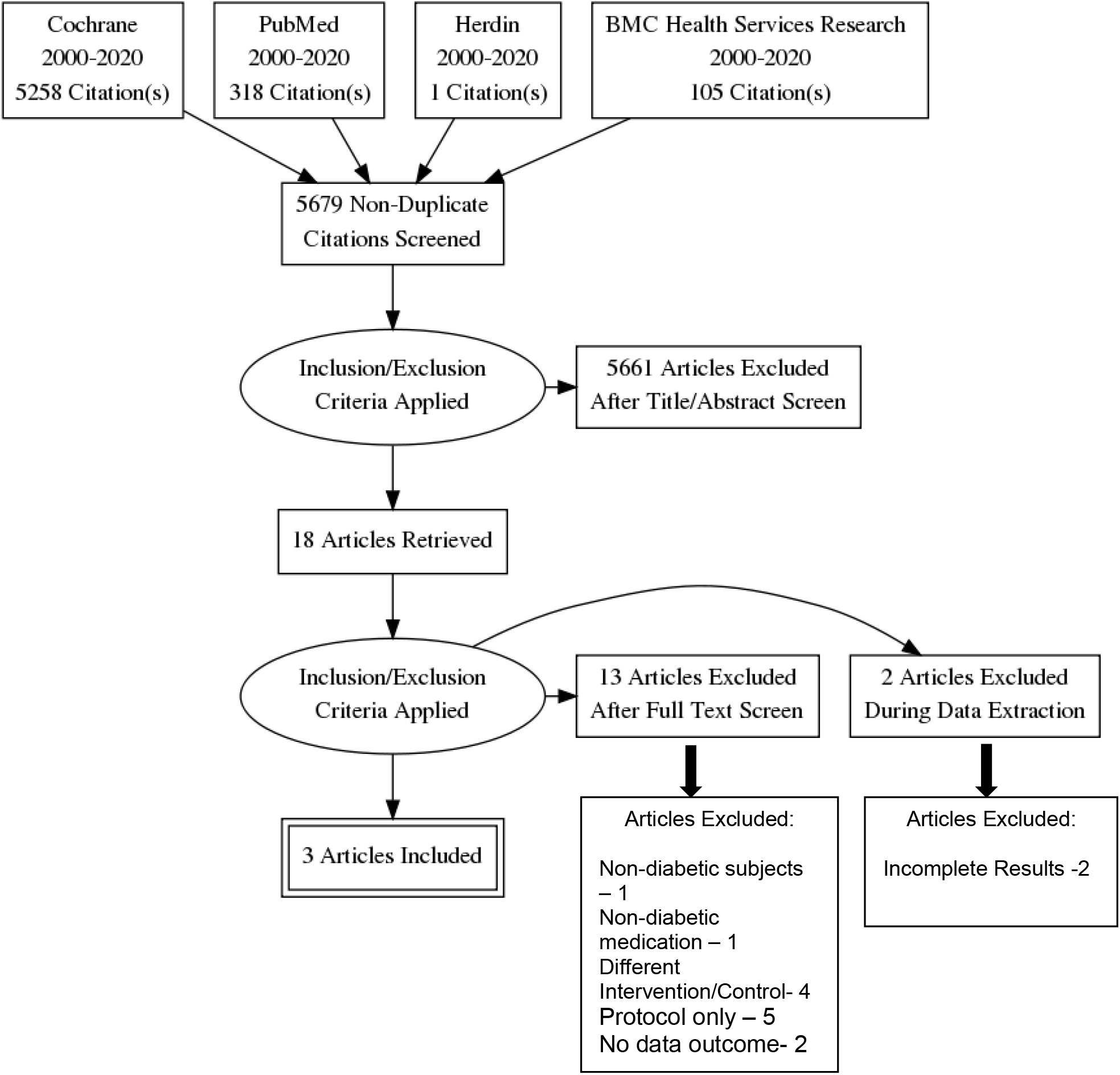
PRISMA Flow Diagram.

### Results

A total of 5679 unique citations were identified through the electronic databases. After title and abstract screening, 5661 articles were excluded. Eighteen articles were assessed for eligibility. After full text screening, thirteen articles were excluded. Two articles were excluded during data extraction since they lack the outcome to be measured in the meta-analysis. Three articles were finally included in the study for analysis.

**TABLE 1:** CHARACTERISTICS OF THE STUDIES.

| Author, Year | Country | Population | Intervention | Comparator | Outcome | Mean, SD |
| --- | --- | --- | --- | --- | --- | --- |
| Owolabi, 2020 (11) | South Africa | IG: 108<br>CG: 108<br>Total: 216 | received daily educational text messages on diabetes and reminders for 6 months | Usual care | On a scale of 8, the mean medication adherence level for the intervention group was 6.90 (SD±1.34) while that of the control group was 6.87 (SD±1.32) with no statistical difference ($P=.88$). | (from 1-8 score)<br>IG: 6.90 (1.34)<br>CG: 6.87 (1.32) |
| O'Connor, 2014 (12) | USA | IG: 341<br>CG: 316<br>Total:657 | one scripted telephone call from a diabetes educator or clinical pharmacist | Usual care | there was no significant difference between the intervention and control arm | (from MPR)<br>IG: 0.802 (0.22)<br>CG: 0.793 (0.24) |
| Arora, 2013 (13) | USA | IG:64<br>CG:64<br>Total:128 | patients received 2messages (9 AM and 5 PM) delivered to their mobile telephones daily for 6 months. | Usual care | medication adherence, which improved from 4.5 to 5.4 in the TExT-MED group compared with a net decrease of – 0.1 in the controls (D1.1; 95% CI 0.1 to 2.1) | (from 1-8 score)<br>IG: 5.4 (2.5)<br>CG: 5.1 (2.5) |

All participants in the three studies were randomized into intervention and control group. Blinding is not plausible since subjects would consent to receive phone call or text message. It was mentioned in the two studies that imputations of missing data and multiple types of analyses were made to avoid attrition bias due to drop outs. One study on the other hand only mentioned multiple analysis for the primary outcome (improvement in HbA1c) and wasn’t clear if the same was done for the secondary outcome (medication adherence). Treatment allocation was not mentioned in 2 studies and would be considered as high risk.

**Figure 2:**
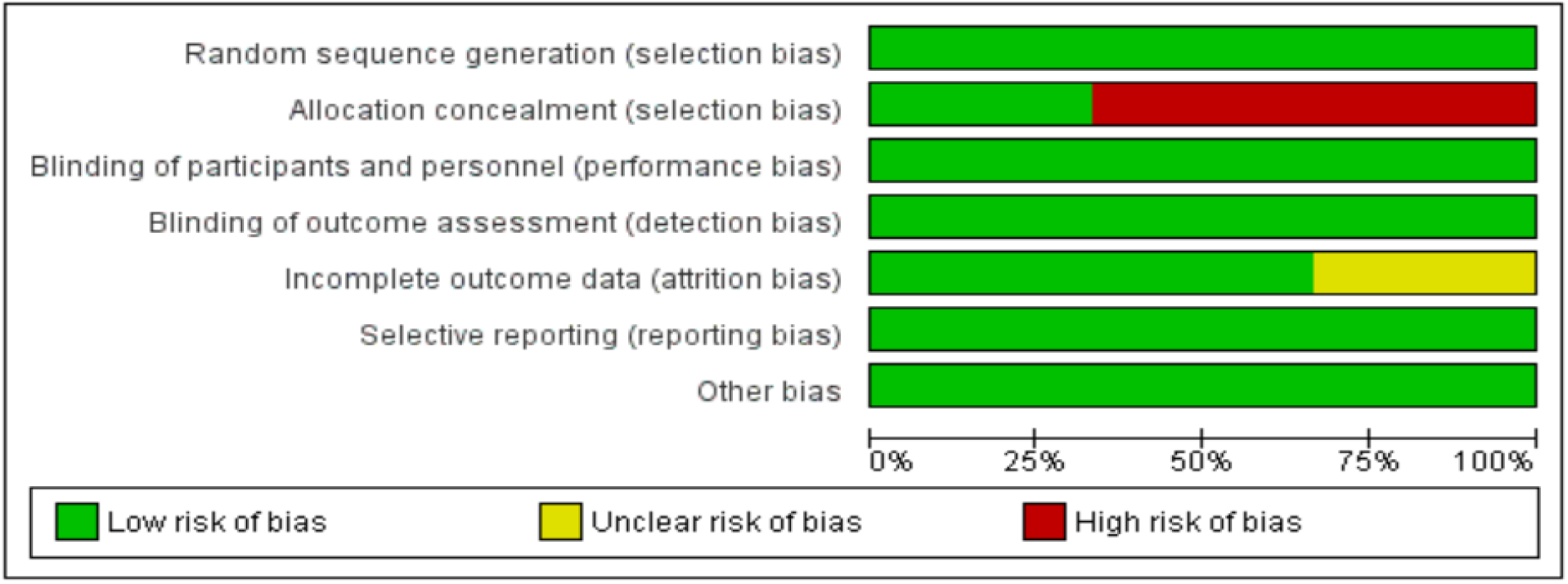
Risk of Bias Graph of Included Trials.

**Figure 3:**
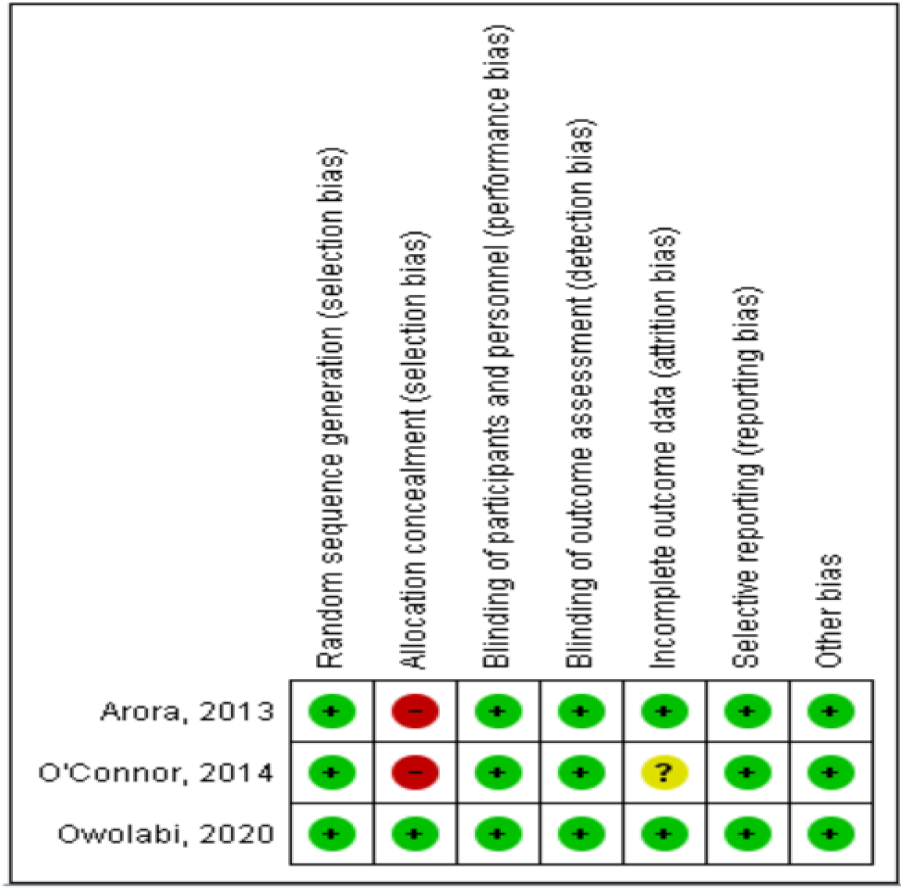
Risk of Bias Summary of Included Trials.

**Figure 4:**
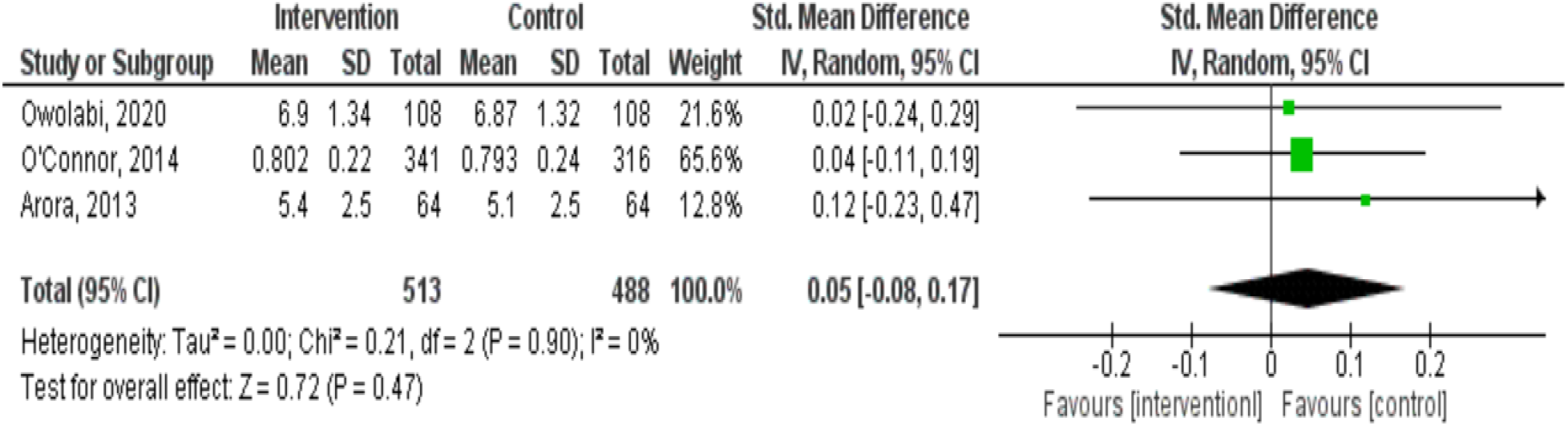
Standard Mean Difference (95% CI) on the effect of phone intervention on medication adherence among diabetic patients.

### Telephone Intervention Efficacy

Three articles were included to investigate the effectiveness of telephone intervention (phone call and text reminders) on the medication adherence among diabetics. Each study reported insignificant finding. The intervention may not have an effect in terms of patients’ adhering to their anti-diabetic medications. The diamond touching the vertical line indicates that the combined results showed to be not statistically significant (SMD: 0.05, 95% CI= -0.08 to 0.17). This may mean that the overall outcome in the telephone intervention group is much the same as in the control group which is the usual care.

I^2^ is 0% which indicates that there is low heterogeneity in the included trials.

## DISCUSSION

This study aims to determine the effectiveness of telephone intervention (phone call and text message reminders) in helping patients adhere to their medications. Three randomized controlled trials were included and pooled effect showed the intervention is not significant in improving medication adherence among diabetic patients.

Owolabi and colleagues (2020) performed a randomized controlled trial among 216 diabetic patients to study the effect of mobile health technology in improving adherence to treatment. Patients (n=108) in the intervention group received daily educational text messages and reminders for 6 months while patients (n=108) in the control group received the usual standard of care only. From 1-8 score, the mean medication adherence level for the intervention group (6.90 (SD±1.34) and control group 6.87 (SD±1.32) show moderate level of adherence but were very much the same scores for both groups. Secondary outcomes included dietary adherence and physical activity adherence. No significant difference between the intervention group and control group were seen on both outcomes. Possible reasons for non-adherence may be due to high cost of healthy diets and lack of motivation to do physical activities. (14)

O’Connor, et al. (2014) evaluated if telephone contact will improve medication adherence, HbA1c, blood pressure and cholesterol level among adults with diabetes who were prescribed with a new class of medication. Intervention group received one scripted telephone call from a diabetes educator or clinical pharmacist and the control group received the usual care. The study started with 1,220 participants assigned to the intervention group and 1,158 assigned to the control group. After 180 days, 657 participants were evaluated for MPR (Medication Possession Ratio): 341 intervention group (mean:0.802, SD: 0.22) and 316 participants for control group (mean:0.793, SD:0.24). According to the authors, the intervention group’s failure to improve adherence, HbA1c, BP and cholesterol level was influenced by many factors and one is due to the low intensity intervention which is only one scripted phone call was used.(12)

Arora, et al. (2013) conducted a study among 128 adult diabetic patients with poorly controlled HbA1C. The intervention group (n=64) received 2 daily text messages for 6 months while the control group (n=64) received the usual care. Using 1-8 score of medication adherence, intervention group score improved from baseline score of 4.5 to 5.4. While on the control group score went down from baseline score of 5.2 to 5.1. There is a trend of slight improvement from the base line but not enough to cause a statistically significant result. Other results include high satisfaction rate for the intervention group (93%) and the lower proportion of patients in the intervention group (35.9%) who used emergency services during the 6-month follow-up period compared with the control group (51.6%). (13)

The non-significant findings of this metanalysis can be seen with the results of the three studies. The intervention group scores on the three trials are very close to the scores of the control group. Limitations of this meta-analysis are the few available trials for analysis and there are two trials included in this study that didn’t mention allocation concealment which may lead to selection bias. Publication bias is unlikely since the insignificant findings were reported. There is low heterogeneity which makes the result safe from inconsistency. The total number of participants exceeded 400 which is considered sufficient and less likely to cause imprecision. With these considerations, this meta-analysis provided moderate strength of evidence.

**TABLE 2:** SUMMARY OF GRADE LEVEL OF EVIDENCE.

| GRADE criteria | Rating (circle one) | Reasons for down- or upgrading | Quality of the evidence (Circle one) |
| --- | --- | --- | --- |
| Study design | RCT (starts as high quality)<br>Non-RCT (starts as low quality) |  | ⊕⊕⊕⊕ High<br>⊕⊕⊕□ Moderate<br>⊕⊕□□ Low<br>⊕□□□ Very Low |
| Risk of Bias | No<br>serious (-1)<br>very serious (-2) | Allocation concealment not mentioned in 2 studies |  |
| Inconsistency (heterogeneity) | No<br>serious (-1)<br>very serious (-2) |  |  |
| Indirectness | No<br>serious (-1)<br>very serious (-2) |  |  |
| Imprecision | No<br>serious (-1)<br>very serious (-2) |  |  |
| Publication Bias | Undetected<br>Strongly suspected (-1) |  |  |
| Other (upgrading factors, circle all that apply) | Large effect (+1 or +2)<br>Dose response (+1 or +2)<br>No Plausible confounding (+1 or +2) |  |  |

## CONCLUSIONS AND RECOMMENDATIONS

In conclusion, this meta-analysis showed that telephone intervention did not result in statistically significant improvement in medication adherence among diabetics. The intervention was no more effective than the usual care.

There are several trials on the effectiveness of telephone intervention on medication adherence but few focused on diabetics. Exploring other interventions for patient adherence such as video calls, mobile applications like viber, messenger and twitter in the Philippine context should be considered in a future study. Other outcomes such as dietary adherence, physical activity adherence, follow-up attendance and quality of life may also be included. Type, number and cost of medications that may influence medication adherence can be further evaluated. To add power to the phone calls or text messages, using the local language or vernacular of the communities may be used for communication. This will be valuable especially to those who cannot speak English or Tagalog. Moreover, all studies included in the systematic review are done prior to the COVID-19 pandemic, and given the health system in general is transitioning to a more virtual approach as part of the adaptive strategies in the new normal, the potential application of mobile communication for treatment adherence will become more popular and a pertinent option for doctors and patients alike

## Data Availability

The authors confirm that the data supporting the findings of this study are available within the article included in the systematic review and its supplementary materials.

## ACKNOWLEDGEMENT

The authors are grateful to Dr Anna Guia Limpoco for her insights, patience, time and reviewing the manuscript.

## OPERATIONAL DEFINITION OF TERMS

1. **Adherence** - “The extent to which a person’ s behavior—taking medication, following a diet, or making healthy lifestyle changes—corresponds with agreed-upon recommendations from a health-care provider”
2. **Diabetes** - is a chronic, metabolic disease characterized by elevated levels of blood sugar which may cause damage to the heart, blood vessels, eyes, kidneys and nerves,
3. **The 8-Item Medication Adherence Scale –** a self-report validated assessment tool that measures non-adherence
4. **Medication Possession Ratio** - used to measure adherence thru refill records

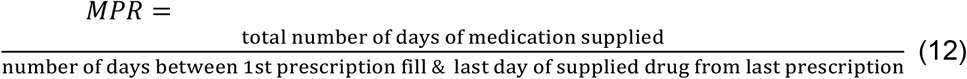

## REFERENCES

1. Brunier A. WHO reveals leading causes of death and disability worldwide: 2000-2019 [Internet]. 9 December 2020. [cited 2021 Jan 18]. Available from: https://www.who.int/news/item/09-12-2020-who-reveals-leading-causes-of-death-and-disability-worldwide-2000-2019

2. LANDICHO-KANAPI MP. DIABETES: Are You At Risk? [Internet]. Philippine Center for Diabetes Education Foundation, Inc. 2016 [cited 2021 Jan 22]. Available from: http://www.pcdef.org/article-of-the-month-may-2016

3. DOH. What can you do to control your blood sugar? | Department of Health website [Internet]. [cited 2021 Jan 22]. Available from: https://doh.gov.ph/faqs/What-can-you-do-to-control-your-blood-sugar

4. WHO. The Impact of Chronic Disease in the Philippines. 2002;12–3.

5. Sabaté E. Adherence to long-term therapies: Evidence for action. Vol. 2, WHO 2003. 2003.

6. Fairman Kathleen Motheral Brenda. Evaluating Medication Adherance: Which Measure Is Right for Your Program? J Manag Care Pharm. 2000;6(6):499–501.

7. Sison G. The Morisky Medication Adherence Scale: An Overview [Internet]. 2018 [cited 2021 Feb 14]. Available from: https://www.pillsy.com/articles/the-morisky-medication-adherence-scale-definition-alternatives-and-overview

8. Jimmy B, Jose J. Patient medication adherence: measures in daily practice. Oman Med J [Internet]. 2011 May [cited 2018 Jul 12];26(3):155–9. Available from: http://www.ncbi.nlm.nih.gov/pubmed/22043406

9. Statista Inc. Smartphone users in the Philippines 2016 | Statista [Internet]. [cited 2017 Apr 1]. Available from: https://www.statista.com/statistics/467186/forecast-of-smartphone-users-in-the-philippines/

10. Kaplan JM. Discharge Follow-Up Phone Calls [Internet]. [cited 2017 Apr 1]. Available from: http://www.jaykaplanmd.com/article-connection-discharge-follow-up/

11. Owolabi EO, Goon D Ter, Ajayi AI. Impact of mobile phone text messaging intervention on adherence among patients with diabetes in a rural setting: A randomized controlled trial [Internet]. Vol. 99, Medicine (United States). Lippincott Williams and Wilkins; 2020 [cited 2021 Jan 19]. Available from: /pmc/articles/PMC7220637/?report=abstract

12. O’Connor PJ, Schmittdiel JA, Pathak RD, Harris RI, Newton KM, Ohnsorg KA, et al. Randomized trial of telephone outreach to improve medication adherence and metabolic control in adults with diabetes. Diabetes Care. 2014 Dec 1;37(12):3317–24.

13. Arora S, Peters AL, Burner E, Lam CN, Menchine M. Trial to examine text message-based mhealth in emergency department patients with diabetes (TExT-MED): A randomized controlled trial. Ann Emerg Med [Internet]. 2014 Jun 1 [cited 2021 Feb 8];63(6):745-754.e6. Available from: http://www.annemergmed.com/article/S0196064413014868/fulltext

14. Owolabi EO, Goon DT AAI. Impact of mobile phone text messaging intervention on adherence among patients with diabetes in a rural setting: A randomized controlled trial. Vol. 99, Medicine (United States). Lippincott Williams and Wilkins; 2020.

